# Classifying post-traumatic stress disorder using the magnetoencephalographic connectome and machine learning

**DOI:** 10.1101/19008037

**Authors:** Jing Zhang, J. Don Richardson, Benjamin T. Dunkley

## Abstract

Given the subjective nature of conventional diagnostic methods for post-traumatic stress disorder (PTSD), an objectively measurable biomarker is highly desirable. Macroscopic neural circuits measured using magnetoencephalography (MEG) has previously been shown to be indicative of the PTSD phenotype and severity. In the present study, we employed a machine learning-based classification framework using MEG neural synchrony to distinguish combat-related PTSD from trauma-exposed controls. Support vector machine (SVM) was used as the core classification algorithm. A recursive random forest feature selection step was directly incorporated in the nested SVM cross validation process (CV-SVM-rRF-FS) for identifying the most important features for PTSD classification. For the five frequency bands tested, the nested CV-SVM-rRF-FS analysis selected the minimum numbers of edges per frequency that could serve as a PTSD signature and be used as the basis for SVM modelling. Many of the selected edges have been reported previously to be core in PTSD pathophysiology, with frequency-specific patterns also observed. Furthermore, the independent partial least squares discriminant analysis suggested low bias in the nested CV-SVM-rRF-FS process. The final SVM models built with selected features showed excellent PTSD classification performance (area-under-curve value up to 0.9). Testament to its robustness when distinguishing individuals from a heavily-traumatised control group, these developments for a classification model for PTSD also provide a comprehensive machine learning-based computational framework for classifying other mental health challenges using MEG connectome profiles.

## Introduction

Post-traumatic stress disorder is a chronic psychological injury that is typically brought about by experiencing or witnessing a life-threatening event (American Psychiatric Association, 1980; Yehuda et al., 2011). The consequences to PTSD include prolonged suffering, distress, impaired quality of life and increased mortality (Kapfhammer, 2014). The disorder is a major neuropsychiatric disorder among military personnel, with up to 17% of Canadian Armed Forces members developing PTSD within the first-year post-deployment (Richardson et al., 2010). The current gold standard for PTSD diagnosis is based on Diagnostic and Statistical Manual of Mental Disorders 5^th^ edition (American Psychiatric Association, 2013). However, these protocols rely heavily on the subjective report of the patients and, given the stigma of a diagnosis in some groups, or difficulty articulating their symptoms, a clear diagnosis can be difficult. As such, an objective diagnosis platform is highly desirable.

One critical step of developing such a framework for PTSD is understanding its psychophysiological and molecular pathology. The underlying neurobiological pathogenesis is increasingly understood within the context of dysfunctional brain circuits (Rauch, Shin & Phelps, 2006). A mechanism that mediates communication and information processing within and between brain circuits is neural oscillations and synchrony (Fries, 2015). Magnetoencephalography (MEG) can image these phenomena non-invasively, and has been used as an effective research tool for exploring the neural activity associated with various neurodegenerative and neuropsychological disorders, including depression, bipolar disorder, mild traumatic brain injury (mTBI) and Alzheimer’s disease (Stam 2010; Vakorin et al., 2016; Alamian et al., 2017; Koelewijn et al., 2019) as well as PTSD-related functional circuitry (Badura-Brack et al., 2018a, 2018b; Dunkley et al., 2014; Mišič et al., 2016). At the group level, neural synchrony can stratify those with PTSD from a heavily-traumatised, but otherwise matched, control group (Misic et al., 2016), with hippocampal synchrony directly related to symptom severity across individuals (Dunkley et al., 2014). This suggests synchrony might be a reliable signature for PTSD identification.

Rapid advancement in artificial intelligence and machine learning have shown promise in brain imaging and computational neuroscience. Various Bayesian inference-based machine learning algorithms have been developed and implemented for neuroimaging signal processing and temporal brain activity prediction (Wu et al., 2016). In translational research and clinical applications, these methods are being actively explored for pre-symptomatic diagnosis, prognostic prediction, and medical intervention effectiveness prediction (Rizk-Jackson et al., 2011). Neurodegenerative and neuropsychological disorders like Huntington’s disease, mTBI and bipolar disorder are among the examples with promising results (Rizk-Jackson et al., 2011; Mitra et al., 2016; Librenza-Garcia et al., 2017).

The objective here was to implement a machine learning classification modelling workflow for delineating individuals with PTSD from trauma-exposed, matched control participants using MEG-derived functional connectomes based on neural synchrony. We developed a comprehensive machine learning pipeline based on support vector machine (SVM) and random forest (RF) algorithms, leveraging their classification modelling and feature selection capabilities, respectively. We recruited combat-related PTSD and trauma-exposed control participants from the Canadian Armed Forces, data that has been published in previous studies (Dunkley et al., 2014; Mišić et al., 2016). This design builds upon our established work and also takes advantage of the similar contexts of traumatic exposure and chronic stress present across participants from serving in a military context, as compared with those from a civilian setting. The present study tests the capacity of machine learning in differentiating PTSD from traumatic-exposure and more generally the potential of this method in distinguishing other mental health challenges.

## Materials and methods

Details on the patient group demographics, data acquisition, and imaging analysis can be found in Dunkley et al., 2014. What follows below in a summary statement. Additional information regarding data collection, processing and machine learning analysis can be found in Supporting Information **S1**. Data acquisition from the 2014 study (Dunkley et al., 2014) was performed with the written informed consent of each individual and under the approval of the Research Ethics Board at the Hospital for Sick Children (SickKids)

### Participants

23 Canadian Armed Forces soldiers diagnosed with PTSD (all male, mean age = 37.4, SD = 6.8, age range 22-48) were recruited and had 5 minutes of eyes open MEG resting state data recorded. Twenty-one trauma-exposed peers (all male, mean age = 33.05, SD = 5.26, age range 18-45) who did not develop PTSD were recruited as a control group.

Inclusion criteria for the PTSD group were: a diagnosis of combat-related PTSD; PTSD symptoms were present from 1-4 years prior to participation in the study; they were engaged in regular mental health follow-up; they had moderate or greater severity on the PTSD check list (PCL>50). All participants in the PTSD group were recruited from one of the Canadian Armed Forces (CAF) Operational and Trauma Stress Support Centres (OTSSC), which are centres of excellence for the diagnosis and treatment of trauma-related mental health injuries. Additional inclusion criteria applied to both groups included: no history of a traumatic brain injury (TBI), screened by a psychiatrist through a review of their electronic health record, telephone interview, and administration of the Defence and Veteran’s Brain Injury Centre (DVBIC) 3 item screening tool; English-speaking and able to understand task instructions and give informed consent. Exclusion criteria included ferrous metal inside the body that might be classified as MRI contraindications or items that might interfere with MEG data acquisition; presence of implanted medical devices; seizures or other neurological disorders, or active substance abuse; certain ongoing medications (anticonvulsants, benzodiazepines, and/or GABA antagonists) known to directly or significantly influence electroencephalographic (EEG) findings. As this was a naturalistic sample, however, all PTSD patients were on evidenced-based psychotropic medication(s), such as selective serotonin reuptake inhibitors (SSRIs), serotonin-norepinephrine reuptake inhibitors (SNRIs) and Prazosin.

### Magnetoencephalography

The details of MEG acquisition and analyses can be found in Dunkley et al., 2014; briefly, we acquired 151 channel MEG on a CTF system at the Hospital for Sick Children. MEG data were coregistered with an anatomical T1 MRI, and a beamformer was used to recover time series from 90 regions of the Automated Anatomical Labelling atlas (AAL) (Tzourio-Mazoyer et al., 2002). The weighted phase lag index (wPLI) was used to determine all pairwise combinations of seed synchrony (Vinck et al., 2011), with wPLI varying between 0 and 1, and used as the edge weight in the matrix. We tested canonical frequency ranges, included Theta (4-7 Hz), Alpha (8-14 Hz), Beta (15-30 Hz), Low Gamma (or L. Gamma, 30-80 Hz) and High Gamma (or H. Gamma, 80-150 Hz). Evaluating multiple frequency ranges allowed us to test the relative performance between the bands, predicting that those which showed the largest group differences previously would provide the greatest accuracy in delineating individual cases here.

### Machine learning downstream data analysis

A visual representation of the overall workflow for the downstream analysis can be viewed in **Fig. 1**. The following core techniques are featured in the method: unsupervised clustering analyses with hierarchical clustering and principal component analysis (PCA), univariate statistical analysis, SVM-centric machine learning analysis, as well as partial least squares discriminant analysis (PLS-DA). Independent from the upcoming machine learning analysis, a univariate analysis step was used to examine the group differences and identify the functional edges with statistically significant changes in connectivity. Data with only significant edges then subject to a machine learning feature selection and modelling process to identify edges essential for classification and subsequently to build a classification model (**Fig. 2**). Similar strategy has been reported elsewhere (Busac et al., 2008). Generally, based on SVM modelling and the previously described recursive RF feature selection (rRF-FS) process (Zhang et al., 2016), a nested cross-validation (CV) framework was used for feature selection (CV-SVM-rRF-FS). Subsequently, data with the selected edges were used for the final SVM classification modelling step. Moreover, PLS-DA was conducted as an independent verification algorithm for the machine learning feature selection and modelling analysis.

**Figure 1.**
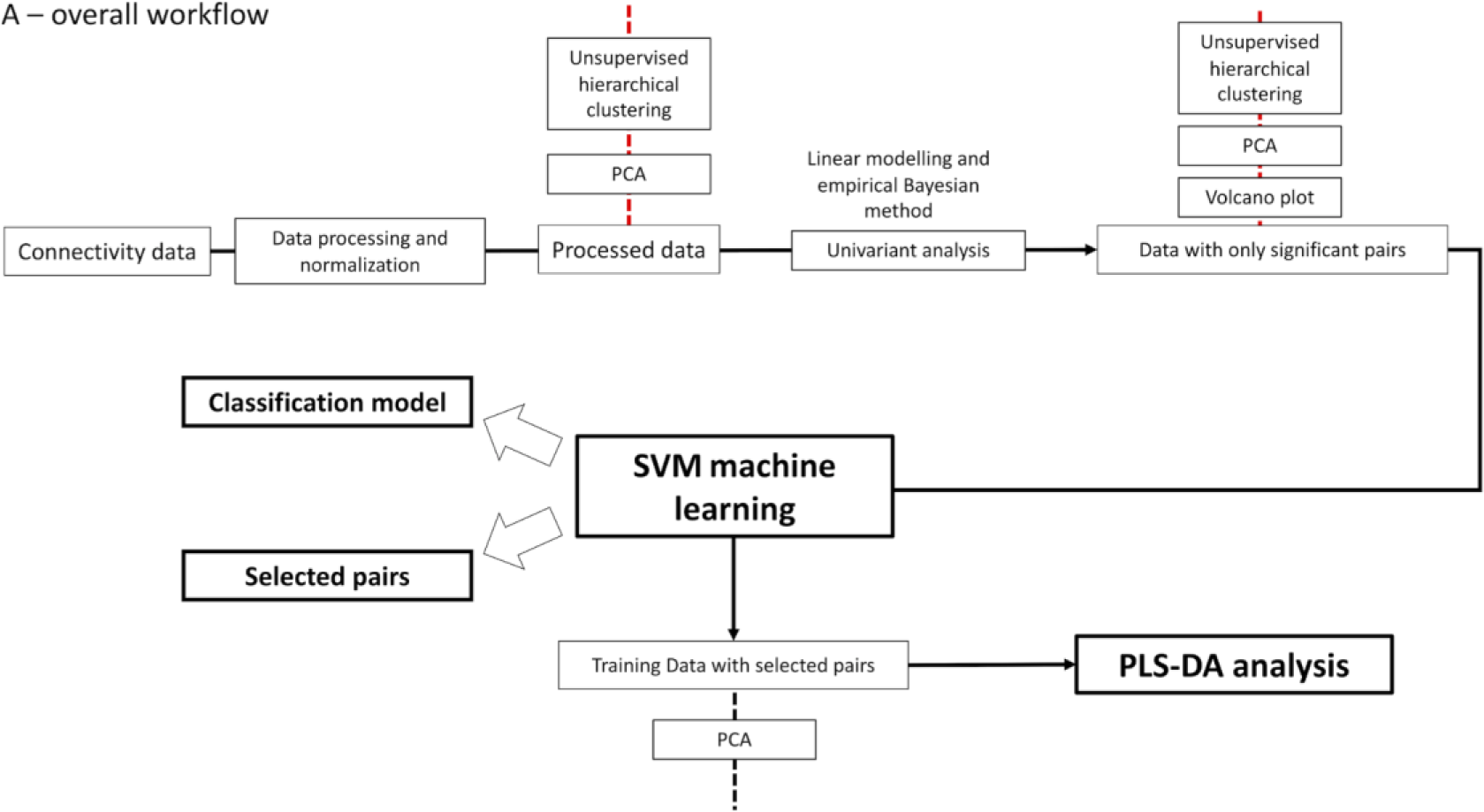
Flowchart for the overall process of machine learning MEG synchrony discovery framework.

**Figure 2.**
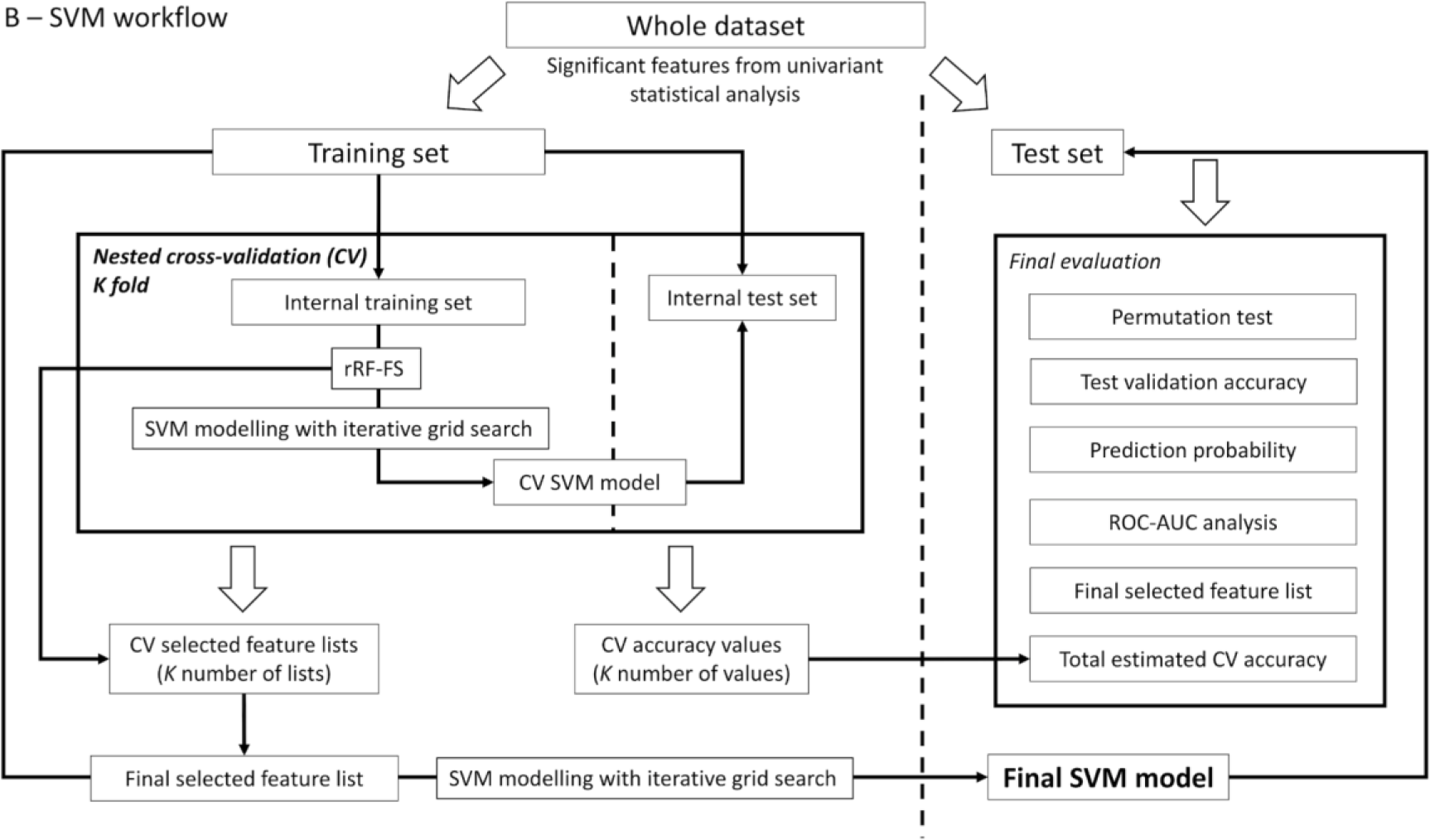
Flowchart for the workflow of the SVM modelling analysis.

Unsupervised clustering analyses were used at various points of the downstream analysis. Specifically, hierarchical clustering was used to explore the grouping pattern in synchrony between the participant groups, as well as between the edges; PCA was used to confirm the functional profile grouping for PTSD and control groups, as well as evaluate data complexity.

The entire downstream data analysis and visualization pipelines were carried out using our custom developed R packages via UNIX Bash scripting. Detailed description of each step in the workflow and software tools used can be found in Supporting Information **S1**.

## Results

### Univariate analysis

A summary of the univariate analysis can be viewed in **Table 1**. The complete univariate analysis along with clustering analysis results are included in Supporting Information **S1** and **Table S1**. Hierarchical clustering was conducted on the data with only the significant edges to assess the clustering pattern upon univariate analysis feature reduction (**Figs 3A, 3B and S3**). In general, reduced data showed improved group clustering results for the five frequency bands. However, we also observed varying results corresponding to specific frequency bands. Regarding the Alpha band (**Fig. 3A**), the clustering result grouped seven participants from the control group into a major cluster, with the second major cluster containing the rest. Within the second major cluster, most PTSD participants were grouped together. For H. Gamma band, with an improved grouping pattern (**Fig. 3B**), the control group exhibited a higher-level of variance than the PTSD group, where six control participants where included in the PTSD cluster. Moreover, the reduced profile in the Theta band managed to mostly separate the PTSD participants from control **(Fig. S3A**). As seen in **Fig. S3B**, the Beta band also showed substantially improved PTSD and control group clustering with the reduced data where only two PTSD participants were placed in the control group. For L. Gamma band (**Fig. S3C**), although two clusters were identified mostly according to the participant groups, the first major cluster only included five control participants, with the rest clustered with the PTSD group to form the second major cluster.

**Table 1.** Feature reduction results summary.

| Frequency band | Significant edges | Increase | Decrease | rRF-FS selected edges | Increase | Decrease |
| --- | --- | --- | --- | --- | --- | --- |
| Theta | 30 | 15 | 15 | 11 | 1 | 10 |
| Alpha | 40 | 22 | 18 | 14 | 9 | 5 |
| Beta | 49 | 30 | 19 | 20 | 10 | 10 |
| L.Gamma | 22 | 7 | 15 | 12 | 4 | 8 |
| H.Gamma | 59 | 26 | 33 | 19 | 9 | 10 |

**Figure 3.**
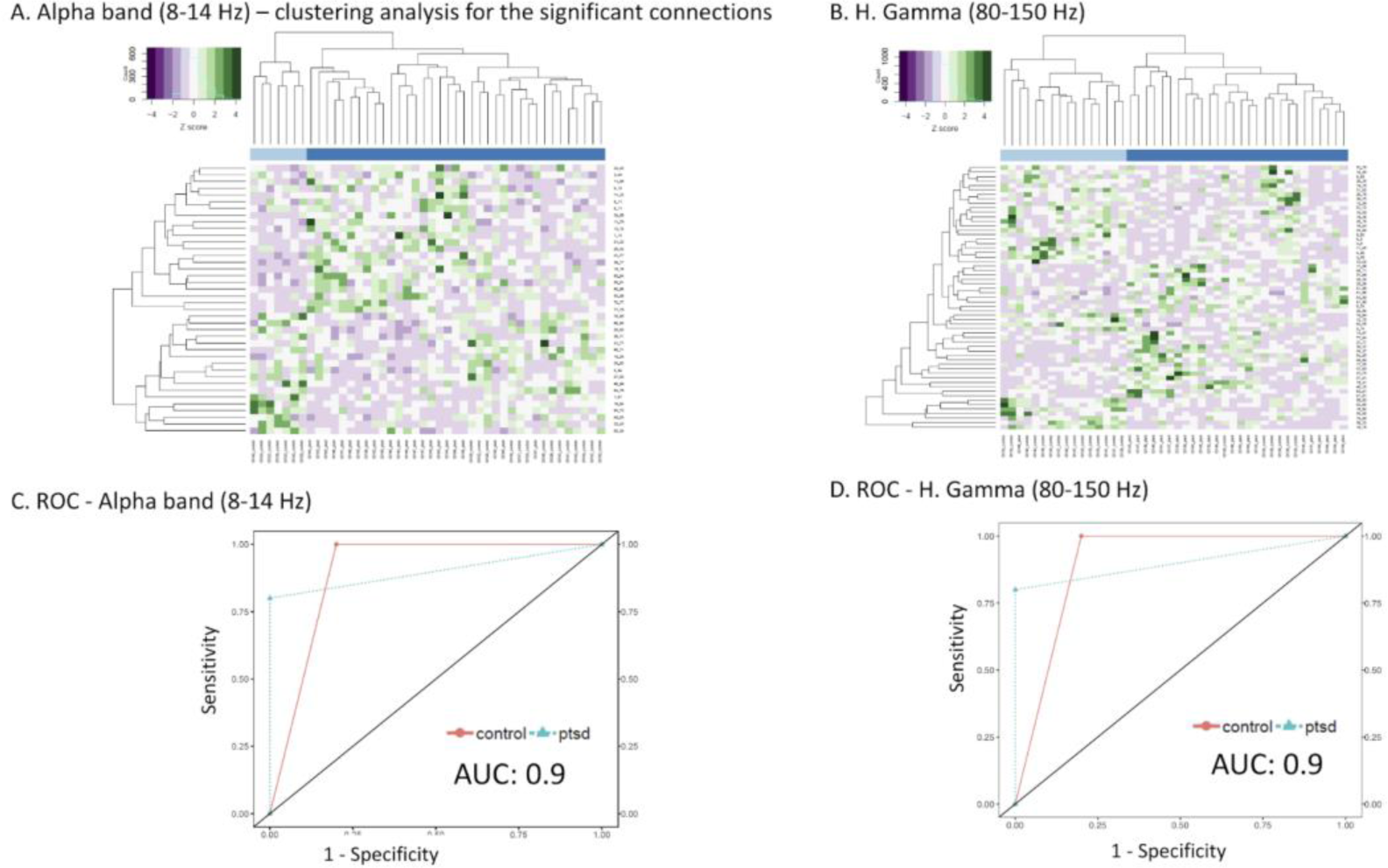
Heatmaps for hierarchical clustering analysis results using only the significant edges as well as ROC-AUC results for the Alpha and H. Gamma bands. For heatmaps, dendrograms show the clusters for participants (columns) and the edges (rows), and z score was plotted. For ROC, AUC values are shown on the plot. (A) Alpha band heatmap, (B) H. Gamma band heaptmap, (C) Alpha band ROC, and (D) H. Gamma band ROC.

### Support vector machine analysis

Using feature-reduced data with only the significant edges identified in the univariate analysis, nested CV-SVM-rRF-FS and classification modelling was carried out. SVM models were trained with the randomly partitioned training set and based on the results from the nested CV featuring rRF-FS. The final SVM models were evaluated by CV step during the modelling process before assessed by the external test set (from the initial random data partitioning step). ROC-AUC analysis with the external test set and permutation test with randomized sample label for training set were used (**Figs. 3C, 3D, S4, S5**). The permutation test suggested that the prediction accuracy of the original model was higher than all the permutation models, leading to a permutation p value at 0.01 across the five frequency bands tested (**Fig. S5**). A summary of the rRF-FS results can be viewed in **Table 1**; and the full list of nested CV-SVM-rRF-FS selected edges can be viewed in **Table 2**. A complete description for SVM modelling results can be found in Supporting Information S1 and **Table S2**.

**Table 2.**
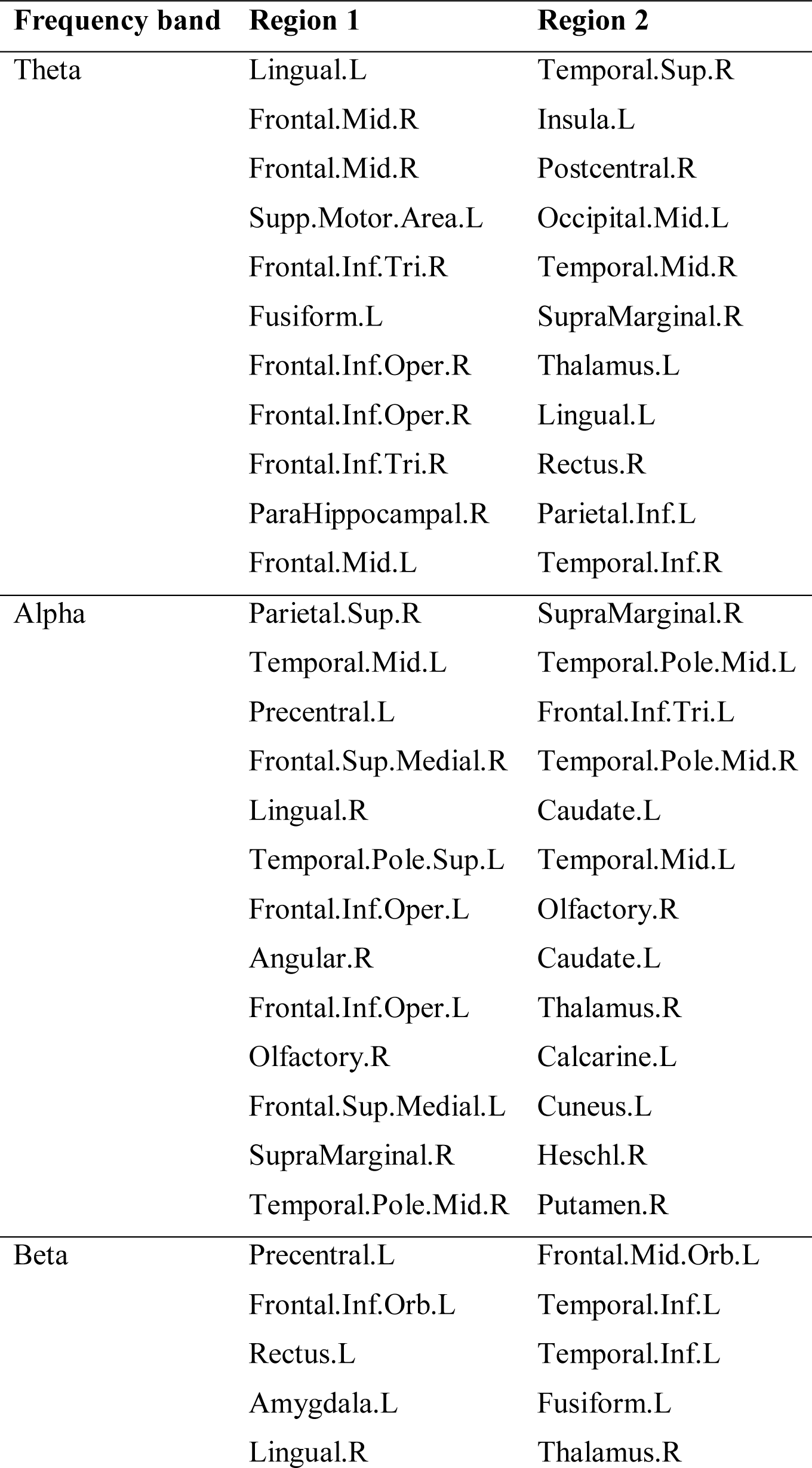
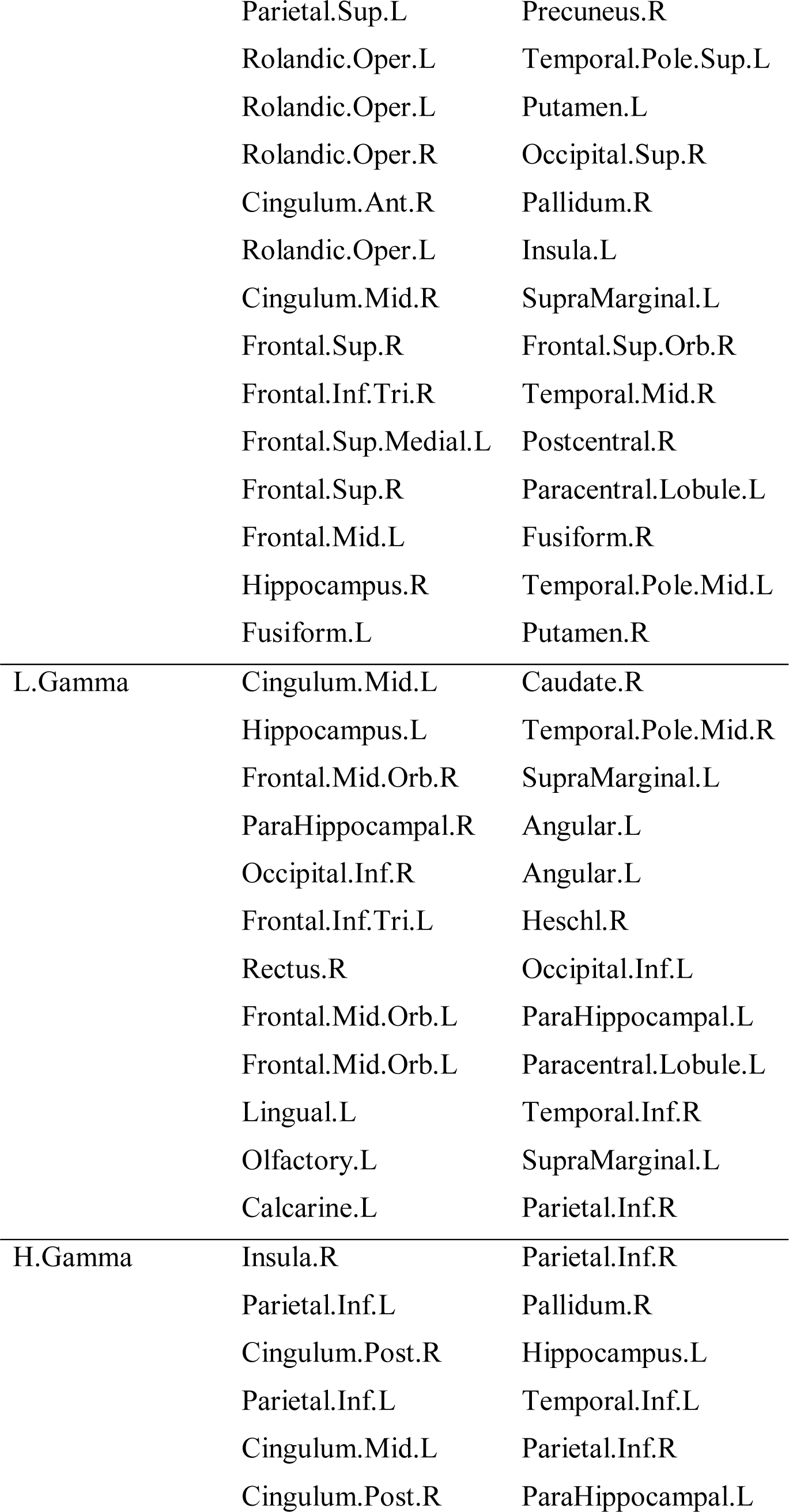
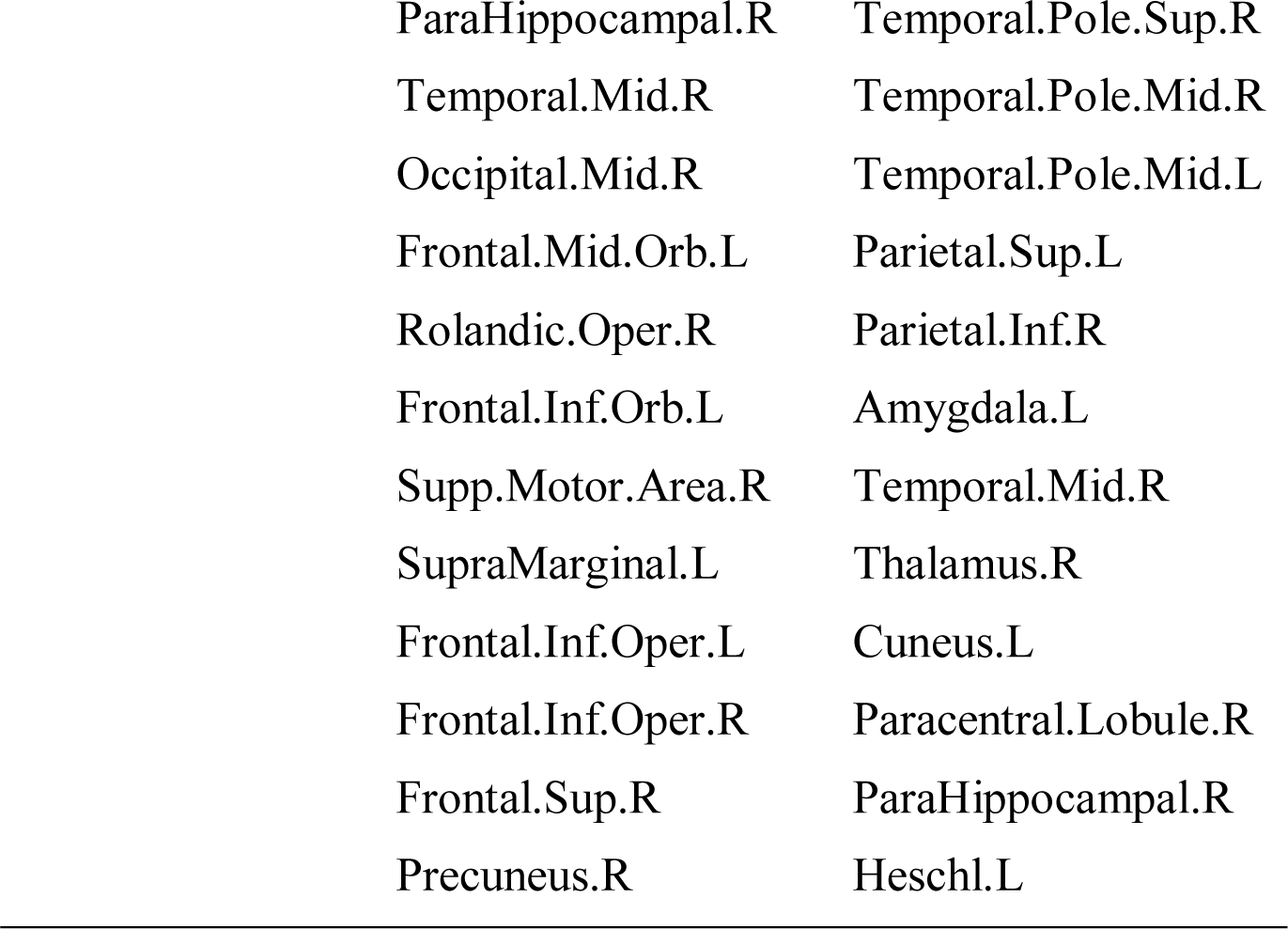
Edges selected by the SVM/rRF-FS process. Abbreviations: inf, inferior; sup, superior; mid, middle; orb, orbital; tri, pars triangularis; oper, operculum; ant, anterior.

| Frequency band | Region 1 | Region 2 |
| --- | --- | --- |
| Theta | Lingual.L | Temporal.Sup.R |
|  | Frontal.Mid.R | Insula.L |
|  | Frontal.Mid.R | Postcentral.R |
|  | Supp.Motor.Area.L | Occipital.Mid.L |
|  | Frontal.Inf.Tri.R | Temporal.Mid.R |
|  | Fusiform.L | SupraMarginal.R |
|  | Frontal.Inf.Oper.R | Thalamus.L |
|  | Frontal.Inf.Oper.R | Lingual.L |
|  | Frontal.Inf.Tri.R | Rectus.R |
|  | ParaHippocampal.R | Parietal.Inf.L |
|  | Frontal.Mid.L | Temporal.Inf.R |
| Alpha | Parietal.Sup.R | SupraMarginal.R |
|  | Temporal.Mid.L | Temporal.Pole.Mid.L |
|  | Precentral.L | Frontal.Inf.Tri.L |
|  | Frontal.Sup.Medial.R | Temporal.Pole.Mid.R |
|  | Lingual.R | Caudate.L |
|  | Temporal.Pole.Sup.L | Temporal.Mid.L |
|  | Frontal.Inf.Oper.L | Olfactory.R |
|  | Angular.R | Caudate.L |
|  | Frontal.Inf.Oper.L | Thalamus.R |
|  | Olfactory.R | Calcarine.L |
|  | Frontal.Sup.Medial.L | Cuneus.L |
|  | SupraMarginal.R | Heschl.R |
|  | Temporal.Pole.Mid.R | Putamen.R |
| Beta | Precentral.L | Frontal.Mid.Orb.L |
|  | Frontal.Inf.Orb.L | Temporal.Inf.L |
|  | Rectus.L | Temporal.Inf.L |
|  | Amygdala.L | Fusiform.L |
|  | Lingual.R | Thalamus.R |
|  | Parietal.Sup.L | Precuneus.R |
|  | Rolandic.Oper.L | Temporal.Pole.Sup.L |
|  | Rolandic.Oper.L | Putamen.L |
|  | Rolandic.Oper.R | Occipital.Sup.R |
|  | Cingulum.Ant.R | Pallidum.R |
|  | Rolandic.Oper.L | Insula.L |
|  | Cingulum.Mid.R | SupraMarginal.L |
|  | Frontal.Sup.R | Frontal.Sup.Orb.R |
|  | Frontal.Inf.Tri.R | Temporal.Mid.R |
|  | Frontal.Sup.Medial.L | Postcentral.R |
|  | Frontal.Sup.R | Paracentral.Lobule.L |
|  | Frontal.Mid.L | Fusiform.R |
|  | Hippocampus.R | Temporal.Pole.Mid.L |
|  | Fusiform.L | Putamen.R |
| L.Gamma | Cingulum.Mid.L | Caudate.R |
|  | Hippocampus.L | Temporal.Pole.Mid.R |
|  | Frontal.Mid.Orb.R | SupraMarginal.L |
|  | ParaHippocampal.R | Angular.L |
|  | Occipital.Inf.R | Angular.L |
|  | Frontal.Inf.Tri.L | Heschl.R |
|  | Rectus.R | Occipital.Inf.L |
|  | Frontal.Mid.Orb.L | ParaHippocampal.L |
|  | Frontal.Mid.Orb.L | Paracentral.Lobule.L |
|  | Lingual.L | Temporal.Inf.R |
|  | Olfactory.L | SupraMarginal.L |
|  | Calcarine.L | Parietal.Inf.R |
| H.Gamma | Insula.R | Parietal.Inf.R |
|  | Parietal.Inf.L | Pallidum.R |
|  | Cingulum.Post.R | Hippocampus.L |
|  | Parietal.Inf.L | Temporal.Inf.L |
|  | Cingulum.Mid.L | Parietal.Inf.R |
|  | Cingulum.Post.R | ParaHippocampal.L |

|  |  |
| --- | --- |
| ParaHippocampal.R | Temporal.Pole.Sup.R |
| Temporal.Mid.R | Temporal.Pole.Mid.R |
| Occipital.Mid.R | Temporal.Pole.Mid.L |
| Frontal.Mid.Orb.L | Parietal.Sup.L |
| Rolandic.Oper.R | Parietal.Inf.R |
| Frontal.Inf.Orb.L | Amygdala.L |
| Supp.Motor.Area.R | Temporal.Mid.R |
| SupraMarginal.L | Thalamus.R |
| Frontal.Inf.Oper.L | Cuneus.L |
| Frontal.Inf.Oper.R | Paracentral.Lobule.R |
| Frontal.Sup.R | ParaHippocampal.R |
| Precuneus.R | Heschl.L |

Here we use Alpha and H. Gamma as examples to exhibit the FS and SVM modelling results. For Alpha, the rRF-FS step nested CV process identified 14 edges as the most relevant features for PTSD-control stratification, including edges between the right superior parietal lobe and supramarginal gyrus, left middle temporal gyrus and middle temporal pole, as well as left precentral gyrus and inferior frontal gyrus pars triangularis. The final Alpha band SVM model was then trained with 31 support vectors with an internal CV accuracy of 0.94. Upon evaluating with external testing data, the AUC value for the Alpha band was 0.9 (**Fig. 3C**). For H. Gamma, 19 edges were selected by the rRF-FS step as the most important features, including edges involving the left amygdala, left hippocampus and thalamus. The nested CV also led to a nested CV accuracy of 0.78 ± 0.06. For the final model, 36 support vectors were used, leading to an internal CV accuracy at 0.94. The AUC value was determined at 0.9 for H. Gamma band (**Fig. 3D**).

### Principal component analysis

Serving as an unsupervised clustering and data complexity assessment tool, PCA was conducted at various points of the downstream data analysis for the five frequency bands tested. The results can be viewed in **Figs. 4** and **S6**. For all five frequency bands, when using all edges, PCA failed to separate the PTSD group from the control participants, whereas the feature-reduced datasets showed substantially improved group clustering. Due to the reduced data dimensionality for the feature-reduced data sets, the data complexity also decreased considerably, which is demonstrated by the increase of the percentage variance explained by the first two PCs in the PCA results.

**Figure 4.**
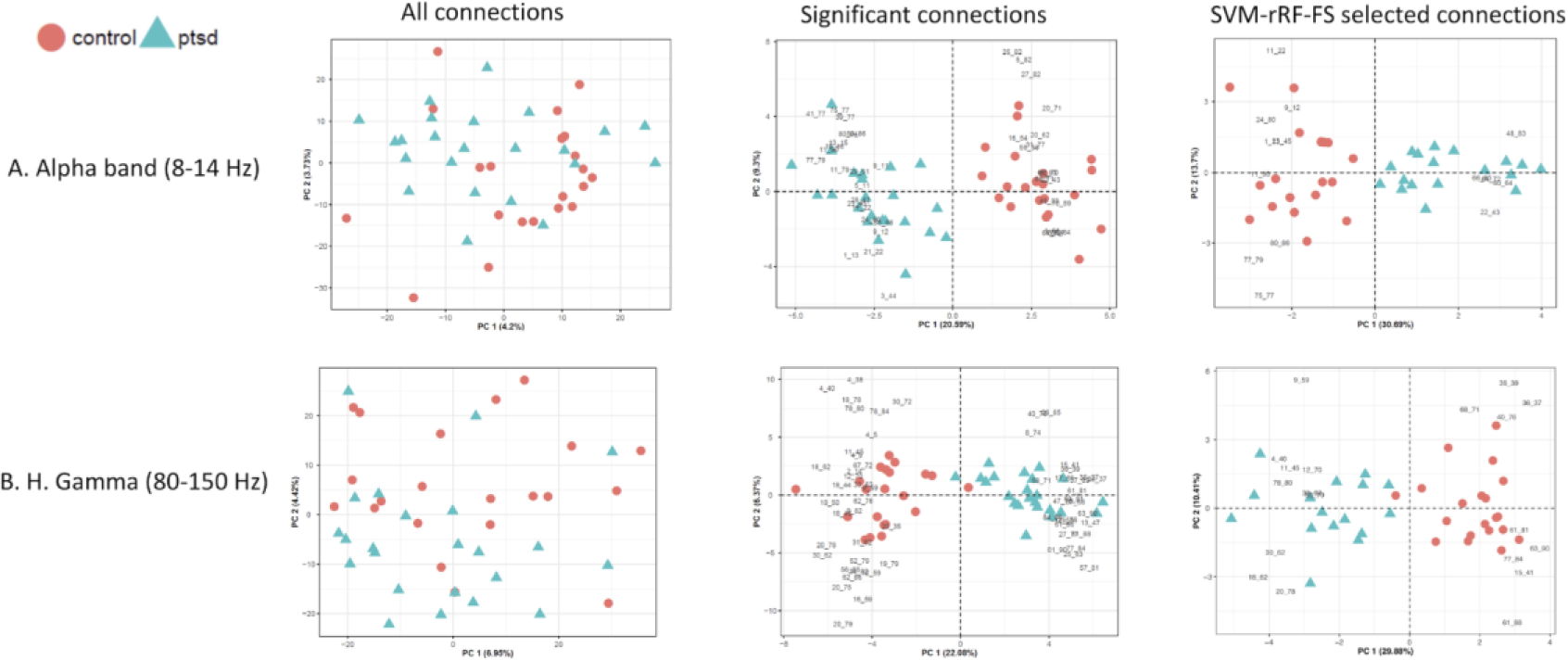
Score plots and biplots (i.e. score plot and loading plot) showing PCA result. For biplot, the loading plots exhibit the contribution of the edges to clustering pattern. Left column: Score plots for PCA results from all the edges; middle column: biplots for PCA results from feature-reduced data with only the significant edges; right column: biplots for PCA results from data with nested CV-SVM-rRF-FS selected edges. (A) Alpha band and (B) H. Gamma band.

### Partial least squares discriminant analysis

PLS-DA results can be viewed in Supporting Information **S1, S2**, as well as **Figs. 5, S7, S8**.

**Figure 5.**
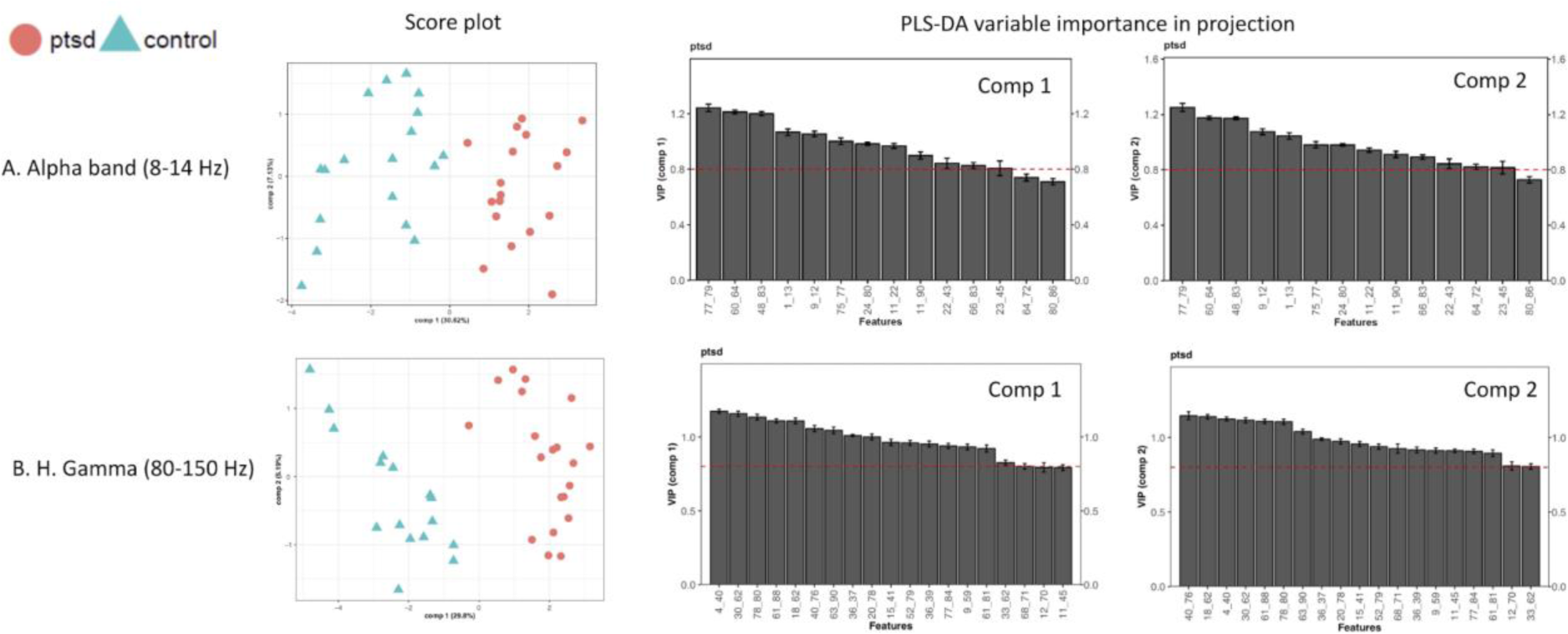
Score plots and VIP plots showing PLS-DA results. Left column: PLS-DA score plots showing the supervised clustering pattern on both components upon PLS-DA modelling using the nested CV-SVM-rRF-FS selected edges; right column: PLS-DA VIP scores for both model components for all the nested CV-SVM-rRF-FS selected edges, with the horizontal dashed line indicating the importance threshold (0.8), and the codes on the x-axis representing the edges. (A) Alpha band and (B) H. Gamma band.

## Discussion

Due to the subjective nature of PTSD diagnostics, the overlap of PTSD symptoms with other disorders (concussion, for example; Garber, Rusu & Zamorski, 2014), and the high comorbidity of other diseased states, including anxiety and depression (Vun et al., 2018), an objectively measurable signature for diagnosing PTSD is desirable. Such a platform may work in concert with the conventional interview and questionnaire-based PTSD diagnostic methods for increased accuracy and facilitating individualized medicine. Given the complexity and dynamic repertoire of neural activity, additional qualifications for an optimal fingerprint include being non-invasive, neurobiologically-informed, as well as high-throughput – this can be achieved by leveraging functional neuroimaging in combination with machine learning and artificial intelligence-inspired approaches.

We considered macroscopic neural circuits based on MEG synchrony as one fingerprint source. Indeed, MEG has been used to understand human neurophysiology, including a wide range of neurodegenerative and neuropsychological disorders, such as mTBI, Alzheimer’s disease, depression and bipolar disorder (Vakorin et al., 2016; Alamian et al., 2017; Koelewijn et al., 2019). Neural oscillations recovered using MEG are known to be dysfunctional in PTSD and correlate with primary symptoms and secondary complaints in the disorder (Kolassa et al., 2007; Dunkley et al., 2014; Mišič et al., 2016; Badura-Brack et al., 2018a, 2018b). Building on this, we developed a comprehensive machine learning-based pipeline for downstream mining and modelling of MEG data. We applied our pipeline to a dataset with over 4000 unique edges, across a number of neurophysiological frequency ranges that are used for multiplexed communication in the brain. It is also worth noting that the current study featured a traumatized control group that had experienced similar combat-related stress as the PTSD cohort, which speaks to the robustness of our machine learning classification framework.

First, unsupervised hierarchical clustering analysis conducted on all the edges showed that the complete functional profiles failed to exhibit any grouping patterns across any of the frequency bands. Moreover, the PCA results on the full dataset with all edges were found to be in complete agreement with the hierarchical clustering analysis, where the score plot exhibited substantial level of overlap among participants from the two groups. These results are at least consistent with the similar life experience the participants with PTSD and control participants had in their military training, deployment and experience of chronic stress and acute trauma during frontline deployment - it is not surprising that the groups possess similar superficial functional profiles in neural activity indexed by synchrony.

Severing as an initial feature reduction method, the univariate analysis substantially reduced the data features for all the frequency bands, with and average number of remaining features at 40±15 (mean±SD), a hundred-fold reduction from the original 4005 edges. The number of significant edges fluctuated according to frequency band, showing frequency-specific patterns. For example, L. Gamma band exhibited the least amount of significant edges, suggesting that the PTSD functional profiles might contain more individual variance across the two participant groups. H. Gamma included the largest number of significant edges, consistent with our previous findings where substantial group difference in neural synchrony was identified for the H. Gamma rhythm (Dunkley et al., 2014; Mišič et al., 2016).

Using the feature-reduced data, the hierarchical clustering and PCA results showed drastically improved group clustering patterns. Specifically, hierarchical clustering exhibited almost complete clustering for the two groups in Theta, Beta and L. Gamma. Even though the Alpha and H. Gamma bands failed to exhibit similar results, the clustering analysis still saw a clear trend of grouping the participants by diagnostic label. Despite the similarity of the complete functional profiles when comparing the two groups, our parametric univariate analysis workflow was able to capture the subtle differences between the two groups. Furthermore, the PCA results suggested that, with the feature-reduced dataset, the two groups could be mostly separated on the first PC. Naturally, the data complexity was also greatly reduced for the data with smaller dimensionality. Consistent with the hierarchical clustering results, PCA also exhibited frequency-specific patterns. Moreover, PCA showed the groups were separated on PC1, which explained the most percentage data variance (around 20%), suggesting the patient/control grouping was the most crucial variable differentiating the data.

The SVM machine learning modelling process was conducted for all five frequency bands using the univariate feature-reduced data. Our 10-fold nested CV-SVM-rRF-FS process produced a consensus edge list. This step further reduced the number of edges by more than 50% across all five frequency bands (**Table 1**). Despite the substantial reduction in data dimensionality, clustering performance stayed mostly unchanged from the univariate feature-reduced data, as seen in the PCA results. This indicates that the current machine learning feature selection strategy was capable of effectively reducing the dimensionality of the data while preserving the information necessary to separate PTSD participants from the control group. Moreover, the PLS-DA VIP (variable importance in projection) evaluation independently confirmed the importance of the selected features. These suggests that our feature selection process was subject to minimal bias.

Frequency-specific patterns were observed for the feature selection results. First, the quantity of the selected edges followed a similar trend as the univariate analysis results, where Beta and H. Gamma exhibited the most selected features, whereas the Theta and L. Gamma bands showed the least. Moreover, based on the corresponding univariate analysis results, we evaluated the directionality distribution for the CV-SVM-rRF-FS selected edges. For example, Theta showed decreased synchrony for 10 out of 11 total selected edges when comparing the PTSD group with controls. Interestingly, a previous study of ours revealed an increase in synchrony in PTSD when compared to controls under the same frequency band, but in a task-dependent manner, during a cognitive flexibility protocol (Dunkley et al., 2015). This suggests the repertoire of neurophysiological activity in PTSD in this particular frequency channel is highly dynamic and flexibly modulated by task. Whilst not examined here, it also suggests using task-induced changes in neural synchrony might be ripe for use as features in machine learning classification for mental illness, as this highly dynamic neural activity is essentially untapped in resting state paradigms. In any case, the initial univariate analysis here revealed an equal number of edges with significant increases and decreases in synchrony, whereas, in addition to drastically reducing the number of edges, CV-SVM-rRF-FS selected more edges with decreased connectivity. These confirmed that while the univariate feature reduction identified the edges with statistical significance at the group level, machine learning was able to further select those features that were most important for individual classification – the increase/decrease ratio compared between the two approaches might not always and necessarily be consistent. Machine learning approaches are ideally suited to recover this granularity. Additionally, the L. Gamma results were mostly consistent with previously reported overall decreases in a range of metrics in the gamma frequency band observed from EEG, such as frontal nodal connection strength and communication efficiency (Lee et al., 2014).

The completely data-driven nested CV-SVM-rRF-FS process was able to extract information that is in line with our knowledge of the neurobiology of PTSD. For example, Theta activity involving the right middle frontal gyrus were among the selected edges with decreased synchrony, including those synchronising with the left insula and right postcentral gyrus. For Alpha, edges involving the right superior parietal lobe and right supramarginal gyrus, left middle temporal gyrus and left middle temporal pole, as well as left precentral gyrus and left inferior frontal gyrus pars triangularis were among the top features differentiating PTSD and controls – other studies have previously reported dysfunction involving the superior parietal lobe, middle temporal gyrus, and left precentral gyrus (Sartory et al., 2013). Additionally, decreased synchrony between the amygdala and fusiform was found to be important for PTSD classification, in line with work from Stevens and colleagues where the weakened coupling between the amygdala and the prefrontal regions was also observed in PTSD conditions (Stevens et al., 2013). L. Gamma synchrony between the left hippocampus and right middle temporal pole also proved to be an important feature, consistent with our previous findings where MEG hyper-synchrony was observed at the group-level for PTSD (Dunkley et al., 2014). Additionally, decreased synchrony between the thalamus and lingual gyrus across multiple frequencies might be related to previously reported findings reporting structural changes in these regions in PTSD (Tan et al., 2013). Taken together, PTSD status was identified using our MEG-derived synchrony and our nested CV-SVM-rRF-FS workflow, highlighting its potential as an application in this disorder and other neuropsychiatric disease, as well as a tool for hypothesis-generation and mechanistic exploration in MEG studies more generally.

Ultimately, a final SVM classification model was built using the CV-SVM-rRF-FS selected data. For the five frequency bands tested, the resulted final SVM models were significant in classifying individuals with PTSD against the trauma-exposed controls according to the permutation tests using the training set. Using the independent test data set, the classification performance showed AUC values over 0.8 for all five frequency bands, suggesting excellent classification accuracy. Notably, Alpha and H. Gamma bands exhibited AUC value of 0.9. Such results were consistent with the previous studies where the Alpha and gamma activity were associated with PTSD in EEG (Clancy et al., 2017; Moon et al., 2018). Additionally, the classification capacity of the selected edges was independently tested using PLS-DA modelling. The results suggested that the high classification capability of the selected edges shown with the SVM modelling also manifest by the PLS-DA modelling, thus considered universal regardless of the classification method.

Not withstanding the limitation of a small sample size in the context of machine learning-based data mining, the present study demonstrated the utility of a comprehensive machine learning pipeline for PTSD classification based off MEG-derived signatures. In summary, the univariate analysis successfully reduced the data size and considerably improved group clustering capacity. The subsequent nested CV-SVM-rRF-FS analysis selected the minimal number of biologically-relevant features that could serve as potential PTSD signatures and be used as the basis for SVM modelling. All final SVM models were significant in classification and exhibited high prediction accuracy, seen by the permutation test and ROC-AUC analysis results, respectively. Furthermore, PLS-DA VIP analysis suggested low method-derived bias for the nested CV-SVM-rRF-FS results. Taken together, the current study not only developed a potential neural circuit marker and an associated classification model for PTSD – it should be remembered this was tested against control participants who themselves were heavily traumatised, some with sub-threshold PTSD symptoms - but also described a machine learning-based computational framework for MEG neural circuit fingerprint discovery and development that could potentially be rolled out to myriad other neuropsychiatric disease.

## Data Availability

This data is available upon reasonable request to the corresponding author.

## Acknowledgements

The authors would like to thanks Margot Taylor, Elizabeth Pang, Rakesh Jetly, Pang Shek, Paul Sedge, Marc Lalancette and Amanda Robertson.

Funded by Defence Research and Development Canada (DRDC), Canadian Institute for Military and Veteran Health Research (CIMVHR), and Innovation for Defence Excellence and Security (IDEaS) program.

## Supporting Information

**S1**. Supplementary methods and results

**S2**. PLS-DA results summary

**Table S1**. Complete results for univariate analysis

**Table S2**. CV-SMV-rRF-FS results with univariate analysis stats.

**Fig. S1**. Heatmaps for hierarchical clustering analysis results using all edges for the five frequency bands. Dendrograms show the clusters for participants (columns) and the edges (rows). (A) Theta band, (B) Alpha band, (C) Beta band, (D) L. Gamma band, and (E) H. Gamma band.

**Fig. S2**. The volcano plots show the edges with significant changes in synchonry (red dots). Horizontal dashed line indicates the p value threshold (0.01) while the vertical line divides directionality (i.e. increases or decreases). (A) Theta band, (B) Alpha band, (C) Beta band, (D) L. Gamma band, and (E) H. Gamma band.

**Fig. S3**. Heatmaps for hierarchical clustering analysis results using only the significant edges for the Theta (A), Beta (B) and L. Gamma (C) bands. Z score is plotted for the heatmaps. dendrograms show the clusters for participants (columns) and the edges (rows).

**Fig. S4**. ROC-AUC results for the Theta (A), Beta (B) and (C) L. Gamma bands. AUC values are shown in the ROC plots.

**Fig. S5**. SVM model evaluation using permutation test. Permutation results showing the percentage accuracy of both the final SVM model and the permutation models, with dashed line indicating the final model accuracy level. Numbers on the x-axis are the models, with 0 representing the final SVM model. (A) Theta band, (B) Alpha band, (C) Beta band, (D) L. Gamma band, and (E) H. Gamma band.

**Fig. S6**. Score plots and biplots (i.e. score plot and loading plot) showing PCA result. For biplot, the loading plots exhibit the contribution of the edges to clustering pattern. Left column: Score plots for PCA results from all the edges; middle column: biplots for PCA results from feature-reduced data with only the significant edges; right column: biplots for PCA results from data with nested CV-SVM-rRF-FS selected edges. (A) Theta band, (B) Beta band and (C) L. Gamma band.

**Fig. S7**. Score plots and VIP plots showing PLS-DA results. Left column: PLS-DA score plots showing the supervised clustering pattern on both components upon PLS-DA modelling using the nested CV-SVM-rRF-FS selected edges; right column: PLS-DA VIP scores for both model components for all the nested CV-SVM-rRF-FS selected edges, with the horizontal dashed line indicating the importance threshold (0.8), and the codes on the x-axis representing the edges. (A) Theta band, (B) Beta band and (C) L. Gamma band.

**Fig. S8**. ROC-AUC and permutation for the PLS-DA models. Left column: ROC curve with AUC values for both components. Right column: Permutation test results showing RMSEP (root mean squared error of prediction) values for both final and permutation PLS-DA models for both participant groups; numbers on the x-axis are the models, with 0 representing the final SVM model. (A) Theta band, (B) Alpha band, (C) Beta band, (D) L. Gamma band, and (E) H. Gamma band.

